# The CRAC channel inhibitor Auxora interrupts inflammatory circuits between alveolar macrophages and T cells in patients with viral pneumonia

**DOI:** 10.64898/2026.05.27.26354034

**Authors:** S. Marina Casalino Matsuda, Vijeeth Guggilla, Catherine A. Gao, Kaitlyn E. Demeulenaere, Luisa Cusick, Samuel W. Fenske, Zhan Yu, Ziyan Lu, Suchitra Swaminathan, Rogan A. Grant, Maxwell J. Schleck, Murali Prakriya, Sudarshan Hebbar, Kenneth Stauderman, Helen K. Donnelly, Chiagozie I. Pickens, Luisa Morales-Nebreda, The NU SCRIPT Study Investigators, Richard G. Wunderink, Alexander V. Misharin, Benjamin D. Singer, G.R. Scott Budinger

## Abstract

Viral pneumonia is perpetuated by inflammatory circuits between activated T cells and monocyte-derived alveolar macrophages (MoAM). T cells and macrophages express ORAI1 and STIM1, which form calcium release-activated calcium (CRAC) channels that allow extracellular calcium entry in response to endoplasmic reticulum calcium store depletion. In a randomized, placebo-controlled, multicenter phase 2 trial (CARDEA), Auxora, a CRAC channel inhibitor, reduced all-cause 30-day mortality by 56% in patients with severe SARS-CoV-2 pneumonia. Here, we report a multi-omics analysis of serially collected alveolar samples from unvaccinated patients with severe SARS-CoV-2 pneumonia treated with Auxora versus placebo. We found reductions in plasma levels of the monocyte– and T cell-chemokines, CCL8 and PDGF-AA. Using peripheral blood mononuclear cells (PBMC) from healthy volunteers, we show that Auxora directly targets T cells to inhibit the transcription of *CCL8* and *PDGFA* in monocyte-derived macrophages, supporting a mechanism for its effects and a potential intermediate biomarker of efficacy.

## Main

In previous reports, we analyzed alveolar samples from patients with severe SARS-CoV-2 pneumonia using flow cytometry, bulk and single-cell RNA-sequencing (scRNA-seq), and cytokine profiling^1-3^. From these data, we generated a model to explain the pathobiology of severe SARS-CoV-2 pneumonia: tissue-resident alveolar macrophages infected with SARS-CoV-2 recruit and activate coronavirus-reactive T cells that produce interferon-γ (IFN-γ). IFN-γ further activates tissue-resident alveolar macrophages and drives their apoptosis. Newly recruited monocytes then differentiate into MoAM, which in turn also become infected and activated, creating a self-sustaining inflammatory signaling loop that drives immunopathology^4^. This model was subsequently credentialed by others in a murine model of influenza pneumonia^5^ and validated in a precision-cut human lung slice model^6^.

CRAC channels are required for T cell activation and contribute to macrophage inflammatory responses^7-9^. These channels are activated when STIM1 in the endoplasmic reticulum (ER) is activated in response to ER calcium store depletion. STIM1 then forms a complex with ORAI1, the pore forming unit of the CRAC channel that allows entry of extracellular calcium^10^. Before safe, effective vaccines protecting against severe SARS-CoV-2 pneumonia were available, we and other investigators enrolled patients with COVID-19 requiring hospitalization and supplemental oxygen in a randomized, placebo-controlled, multi-center phase 2 trial of Auxora (CARDEA), which resulted in a 56% reduction in 30-day all-cause mortality^11^. All patients in the trial received corticosteroids, and some received other immunomodulatory therapies for COVID-19. These promising phase 2 results have prompted ongoing trials of Auxora in other critically ill patients.

Concurrent with the CARDEA trial, we designed and enrolled patients with laboratory-confirmed SARS-CoV-2 pneumonia complicated by the acute respiratory distress syndrome in a single-center, placebo-controlled, ascending-dose phase 2 clinical trial to generate pharmacokinetic (PK) and pharmacodynamic (PD) data in critically ill, mechanically ventilated patients (NCT04661540) (**Fig. 1a**). Inclusion and exclusion criteria, study design and enrollment, and patient demographics are found in the online supplement (**Supplementary Fig. 1 and Supplementary Table 1**). SARS-CoV-2 viral titers estimated from the RT-qPCR cycle threshold of BAL fluid did not differ in patients receiving Auxora or placebo (**Fig. 1b**). Mortality and duration of ventilation did not differ in patients who received Auxora or placebo, but the only patients to be discharged alive and off ventilation by day 30 received Auxora (**Fig. 1c**). Plasma levels of Auxora are shown in **Fig. 1d**. BAL fluid levels of Auxora were below the limit of detection in all but 3 samples. We measured plasma levels of 72 cytokines, 55 of which were of sufficient quality for downstream analysis^2^. The levels of CCL8 (aka MIP-2) which is nearly exclusively produced by macrophages and involved in the recruitment of monocytes, and PDGF-AA a mitogen and chemoattractant of mesenchymal cells, were significantly reduced after receiving Auxora but not placebo (**Fig. 1e**).

**Figure 1:**
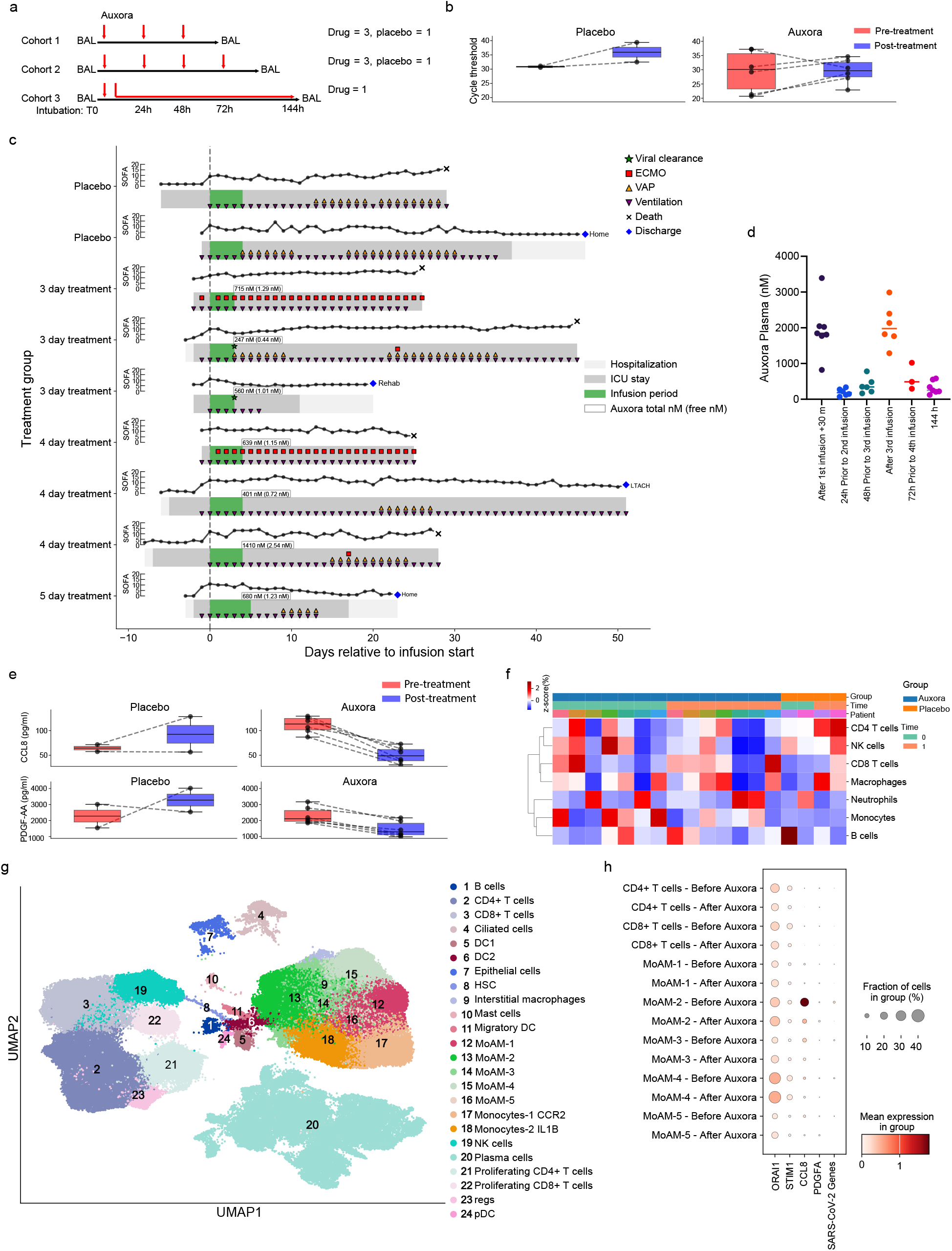
Auxora decreases levels of CCL8 and PDGF-AA in plasma of patients with severe SARS-CoV-2 pneumonia. **a**, Schematic illustrating protocol driven interventions. The number of patients in each cohort and treatment group is indicated. Time 0 is defined by the initial BAL and drug infusion. **b**, Viral load before and after placebo or Auxora infusion estimated by PCR cycle threshold in BAL fluid samples. **c**, Swimmer plots of the ICU course in patients enrolled in the trial. Sequential organ failure assessment (SOFA) score, assessment of viral clearance, initiation of extracorporeal membrane oxygenation (ECMO), ventilator associated pneumonia episode (VAP). **d**, Auxora total concentrations in plasma. **e**, Plasma CCL8 and PDGF-AA levels before and after placebo or Auxora infusion. Significance determined by linear mixed models using a paired design. **f**, Cell abundance measured by flow cytometry analysis of BAL samples. Samples were clustered by Manhattan distance using complete linkage. **g**, Uniform manifold approximation and projection (UMAP) plot showing integrated analysis of BAL cells from patients with severe SARS (n = 9). **h**, Dot plot of *ORAI1, STIM1, CCL8*, PDGF-AA, and SARS-CoV-2 genes, in different cell types before and after Auxora treatment. Differences were not significant based on differential gene expression analysis using DESeq2 (q > 0.05, Wald test with FDR correction). Statistical testing not performed for SARS-CoV-2 genes due to sparse expression.

Auxora treatment did not result in substantial differences in BAL fluid immune cell composition measured by flow cytometry (**Fig. 1f, Supplementary Fig. 2**). For scRNA-seq on BAL fluid cells, we clustered and annotated cells as previously described (**Fig. 1g and Supplementary Figs. 3a**,**b**). Consistent with our flow cytometry results, we did not detect substantial differences in the cellular composition of the BAL fluid after treatment with Auxora (**Supplementary Fig. 3b**). We did not detect differences in gene expression in any cell population as a function of Auxora treatment (**Supplementary Fig. 3c**). *CCL8* expression was limited to alveolar macrophages, and while the differences did not reach statistical significance, *CCL8* were lower in MoAM from patients treated with Auxora compared with their pretreatment baseline (**Fig. 1h**). SARS-CoV-2 transcripts were detected in MoAM at baseline and were nearly absent across all cell types following Auxora treatment. Differential expression testing was not performed for viral genes due to sparse expression and near absence of detectable viral RNA in post-treatment samples (**Fig. 1h**).

**Figure 2:**
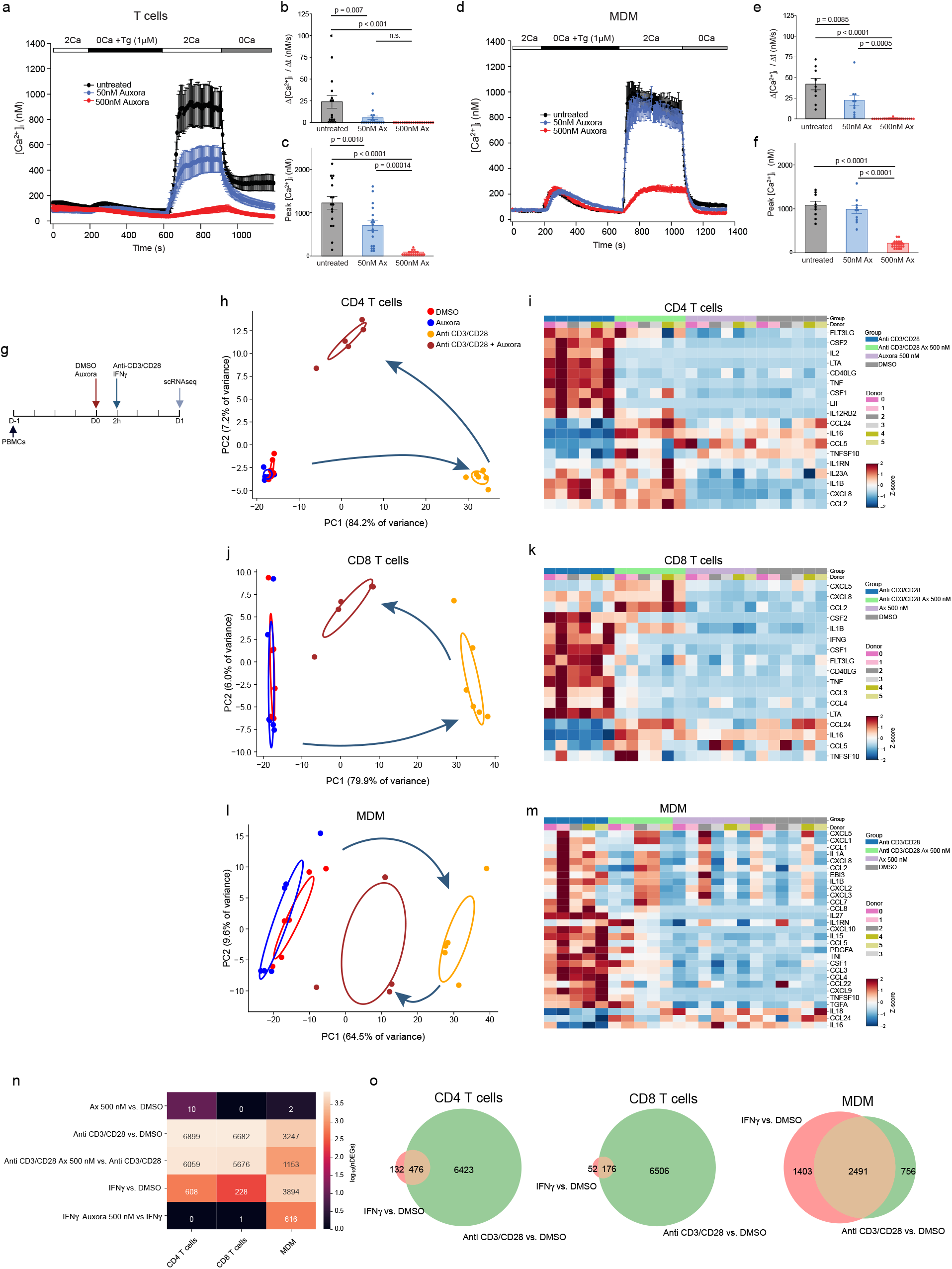
Auxora targets T cells to inhibit T cell-macrophage circuits during SARS-CoV-2 pneumonia. **a-f**, We measured store-operated calcium entry (SOCE) in Fura-2 loaded cells by administering thapsigargin (Tg, 1 μM) in Ca^2+^-free solution followed by the addition of extracellular Ca^2+^ (2 mM) containing media. **a**, Time course of calcium measures in peripheral T cells. **b**, Rate of calcium influx after addition of extracellular Ca^2+^ following store depletion. **c**, Total amount of calcium over baseline entering the cell in the five minutes after switch to Ca^2+^ (2 mM) containing media. **d-f**, same as **a-c** in monocyte-derived macrophages (MDM). PBMCs were isolated from 3 different healthy donors. Bar graphs represent mean ± SEM. T cells, untreated n = 16 cells; 50 nM Auxora n = 18 cells; 500 nM Auxora n = 19 cells. MDM, untreated n = 9 cells; 50 nM Auxora n = 10 cells; 500 nM Auxora n = 22 cells. **g**, Schematic for experiments in PBMCs from 5 different blood donors in **h-o. h-m**, Genes were identified as differentially expressed by a negative binomial generalized linear model using an ANOVA-like test for differential expression between any conditions in the dataset (FDR q < 0.05), Ax=Auxora. **h**, PCA of differentially expressed genes in CD4 T cells. **j**, CD8 T cells **l**, MDM. **i**, Heat map demonstrating the expression of selected genes of interest among the differentially expressed genes in CD4 T cells, **k**, CD8 T cells and **m**, MDM. **n**, Heatmap showing the number of differentially expressed genes for each comparison. **o**, Venn diagrams indicate overlap between differentially expressed genes induced by treatment with anti CD3/CD8 antibodies or recombinant IFN-γ.

Both alveolar macrophages and T cells express *ORAI1* and *STIM1* (**Fig. 1h**). We compared the pharmacodynamic responses of T cells and macrophages to Auxora using PBMC from healthy volunteers after differentiating monocytes into monocyte-derived macrophages (MDM). In T cells, we found that Auxora blocked Ca^+2^ entry in a dose-dependent fashion beginning at a concentration of 50 nM (**Figs. 2a-c**). In contrast, Auxora inhibited CRAC channel-mediated Ca^2+^ entry in MDM at a concentration of 500 nM (**Figs. 2d-f**).

We sought to test whether Auxora inhibits inflammatory circuits between MDM and T cells, and if so, whether it targets T cells, MDM, or both. We cultured PBMC from healthy volunteers for 24 hours, then treated them with Auxora (500 nM) or vehicle, followed 2 hours later by anti-CD3/CD28 antibodies or IFN-γ, and processed them for scRNA-seq 24 hours later (**Fig. 2g; Supplementary Figs. 4a-d**). Auxora did not induce cytotoxicity at this concentration (**Supplementary Fig. 4a**). Principal component analysis (PCA) revealed that treatment with anti-CD3/CD28 antibodies induced transcriptional reprogramming in CD4 and CD8 T cells and MDM that was attenuated by Auxora (**Figs. 2h, j, and l**). We identified large numbers of differentially expressed genes in CD4 and CD8 T cells and MDM treated anti-CD3/CD28 antibodies, a majority of which were affected by Auxora (**Supplementary Fig. 5a**). Genes attenuated by Auxora included those encoding secreted cytokines involved in T cell activation, for example *IL2* and *TNF*, and *IFNG* (**Figs. 2i, k, and m**). GSEA revealed activation of several common inflammatory pathways in T cells and MDMs that were inhibited by Auxora, for example, TNFα signaling via NFκB and IL2 Stat5 signaling (**Supplementary Figs. 5b-c)**.

As MDMs do not express CD3 or CD28, their activation in response to anti-CD3/CD28 antibodies is likely attributable to indirect signals from T cells (**Figs. 2m-o and Supplementary Fig. 4g**). Accordingly, we sought to test whether the inhibitory effects of Auxora on MDM were mediated by its effects on T cells or a direct effect on MDM. We observed no effect of Auxora when we treated MDM with TLR agonists in the absence of T cells (**Supplementary Figs. 6a-b**). We then examined the effect of Auxora on the response of T cells and MDM to recombinant IFN-γ, the primary cytokine released by T cells in the single-cell data (**Supplementary Figs. 4e-g**). Only a small fraction of the genes induced in T cells by anti-CD3/CD28 antibodies were induced by IFN-γ. In contrast, genes induced in MDM treated with IFN-γ substantially overlapped with those induced by anti-CD3/CD28 antibodies and IFN-γ in MDM (**Figs. 2n-o**). These results suggest that Auxora targets T cells to interrupt inflammatory circuits with MoAM, mediated in large part by IFN-γ. Identifying pneumonia endotypes characterized by similar circuits might guide the selection of patients for future clinical trials of Auxora in patients with pneumonia.

## Data Availability

All data produced in the present work are contained in the manuscript

https://github.com/NUPulmonary/Guggilla_Matsuda_Auxora_2026

https://sqlifts.fsm.northwestern.edu/internal/auxora/?ds=pbmc_integrated_0424

https://sqlifts.fsm.northwestern.edu/internal/auxora/?ds=2025-09-30_Auxora_Human

## Acknowledgments

C.A.G. is supported by NIH/NHLBI K23HL169815, a Parker B. Francis Opportunity Award, and an American Thoracic Society Unrestricted Grant. R.G.W. is supported by NIH/NIAID U19 AI135964, R01 AI158530; NHLBI R01 HL149883, P01 HL154998; U01TR003528. B.D.S. is supported by NIH awards R01HL149883, R01HL153122, P01HL154998, P01AG049665, U19AI135964, and U19AI181102. G.R.S.B. is supported by the NIH (U19AI135964, P01AG049665, R01HL147575, P01HL071643, U54AG079754, R01HL145478, R01HL147290, R01HL173940, U19AI181102, and R01HL154686); the US Department of Veterans Affairs (I01CX001777); Simpson Querrey Lung Institute for Translational Sciences. A.V.M. was supported by the NIH (grant nos. U19AI135964, P01AG049665, P01HL154998, P01HL169188, U19AI181102, R01HL153312, R01HL158139, and R01ES034350), and research grants from AbbVie and Merck.

We thank all patients and volunteers who provided their samples and data for this study and their families. This research was supported in part through a generous gift from Kimberly Querrey and Louis A. Simpson. This research was supported by the Simpson Querrey Lung Institute for Translational Science (SQLIFTS), Northwestern University. This research was also supported by the computational resources and staff contributions provided for the Quest high-performance computing facility at Northwestern University, which is jointly supported by the Office of the Provost, the Office for Research and Northwestern University Information Technology. This research was also supported in part through the computational resources and staff contributions provided by the Genomics Compute Cluster, which is jointly supported by the Feinberg School of Medicine, the Center for Genetic Medicine and Feinberg’s Department of Biochemistry and Molecular Genetics, the Office of the Provost, the Office for Research and Northwestern Information Technology. The Genomics Compute Cluster is part of Quest, Northwestern University’s high-performance computing facility, with the purpose of advancing research in genomics. Northwestern University Flow Cytometry Core Facility is supported by the National Cancer Institute Cancer Center support grant (no. P30 CA060553) awarded to the Robert H. Lurie Comprehensive Cancer Center. Sorting was performed on a BD FACS Aria SORP cell sorter purchased with the support of the National Institutes of Health (NIH, grant no. 1S10OD011996-01). Integrative genomic services were performed by the Metabolomics Core Facility at Robert H. Lurie Comprehensive Cancer Center of Northwestern University.

## Author contributions

Conceptualization: S.M.C.M., R.G.W., B.D.S., G.R.S.B., and A.V.M. Methodology: S.M.C.M., V.G., C.A.G., R.G.W., and A.V.M. Software: S.M.C.M., V.G., C.A.G., L.C., S.W.F., M.S. and A.V.M. Validation: S.M.C.M., V.G., C.A.G., R.G.W., and A.V.M. Formal analysis: S.M.C.M, V.G., C.A.G., L.C., S.W.F., R.G.W., and A.V.M. Investigation: S.M.C.M., C.A.G., K.D., Z.Y., Z.L., R.A.G., H.K.D., R.G.W., and A.V.M. Resources: S.M.C.M., V.G., C.A.G., K.D., Z.Y., Z.L., S.S., R.A.G., M.P., S.H., K.S., H.K.D., C.I.P., L.M.-N., R.G.W., B.D.S., G.R.S.B. and A.V.M. Data curation: S.M.C.M, V.G., C.A.G., L.C., Z.Y., Z.L., R.G.W., B.D.S., G.R.S.B. and A.V.M. Writing – all aspects: S.M.C.M., R.G.W., B.D.S., G.R.S.B., and A.V.M. Writing – review & editing: S.M.C.M. Visualization: S.M.C.M. and V.G. Supervision: S.M.C.M., R.G.W., G.R.S.B., B.D.S., and A.V.M. Project administration: S.M.C.M., R.G.W., B.D.S., G.R.S.B., and A.V.M. Funding acquisition: R.G.W., B.D.S. G.R.S.B., and A.V.M.

