## Supplemental figures and table for "The CRAC channel inhibitor Auxora interrupts inflammatory circuits between alveolar macrophages and T cells in patients with viral pneumonia"

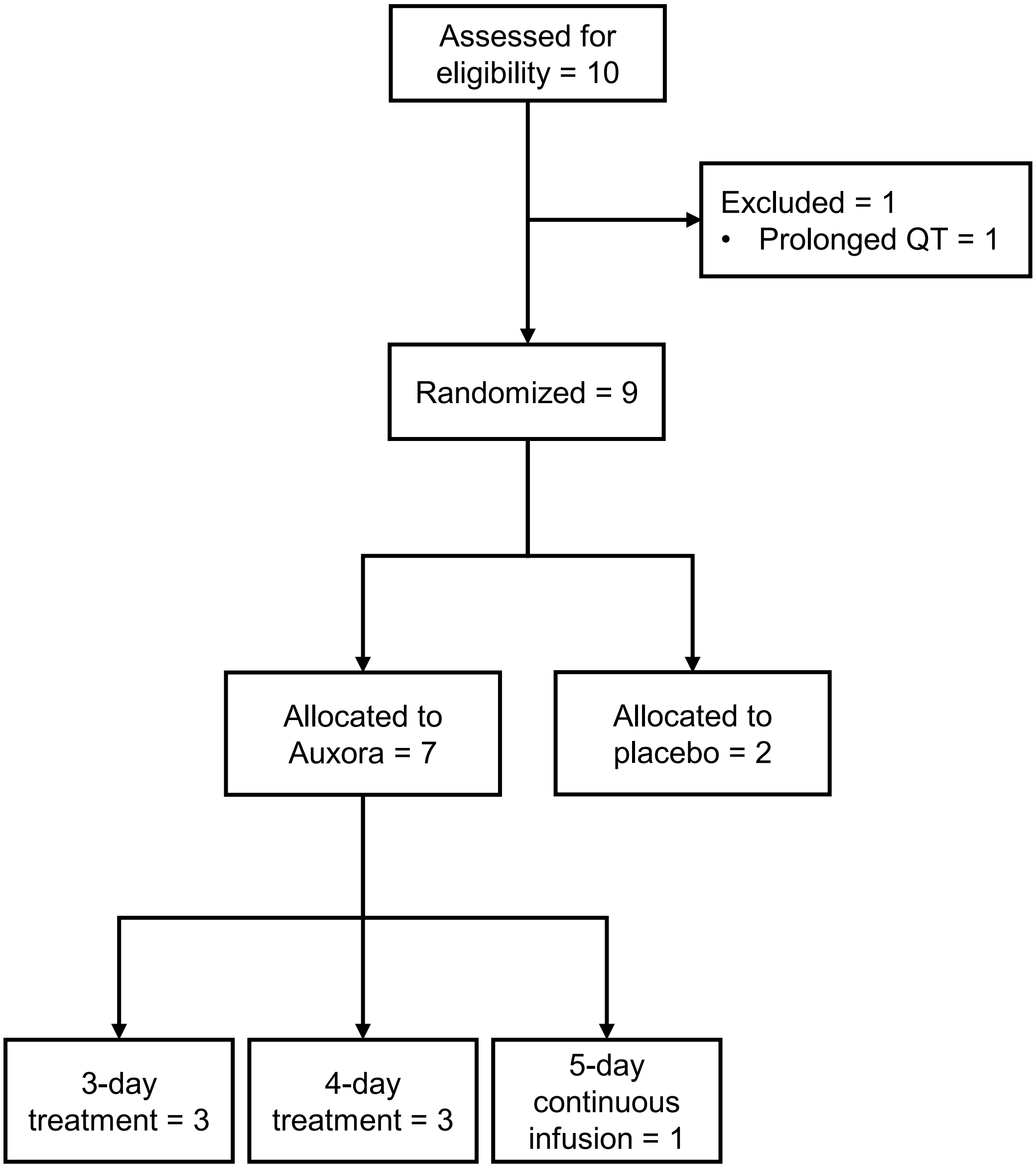


**Supplementary Figure 1: CONSORT diagram of the study.**

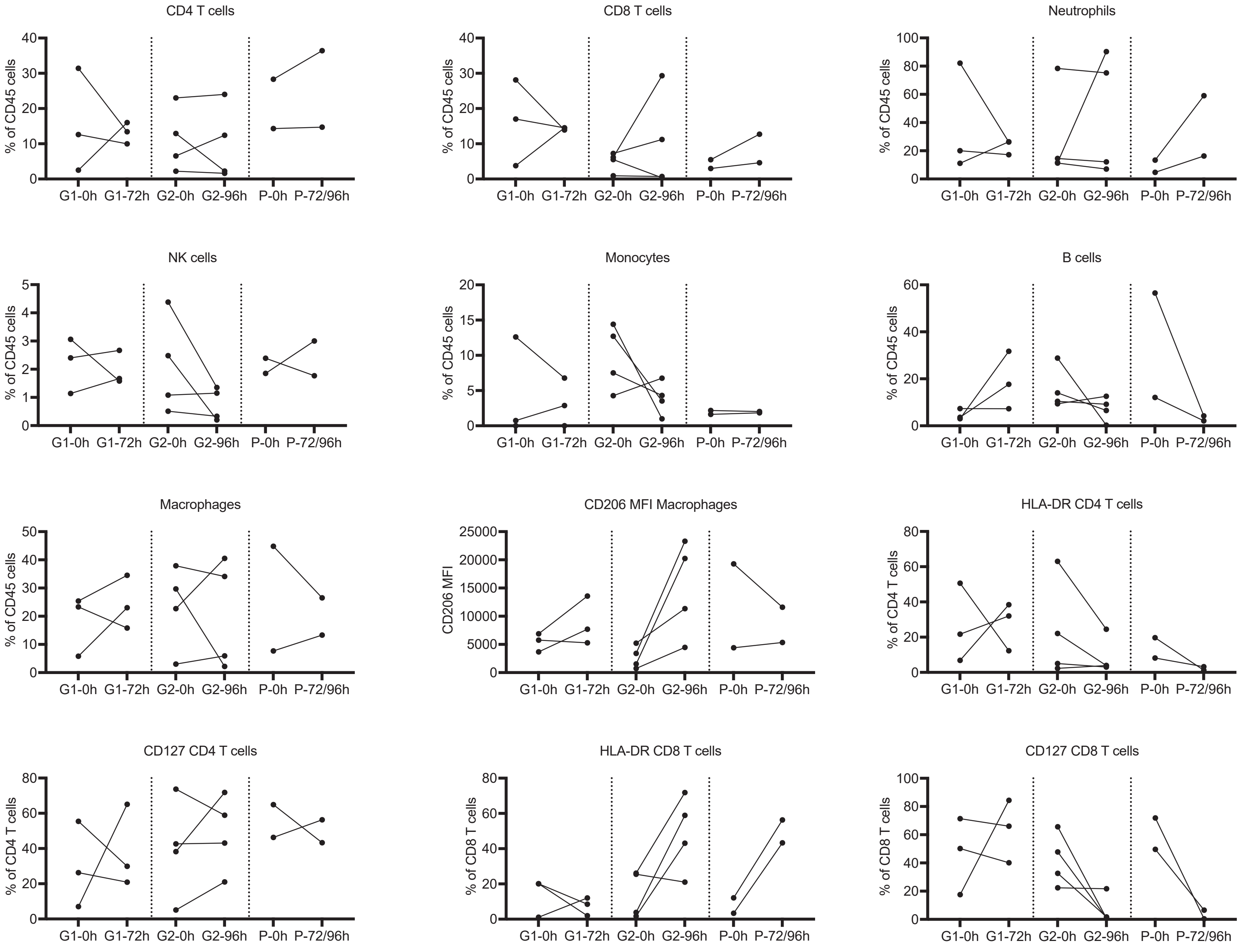


<u>**Supplementary Figure 2:**</u> **Auxora does not affect the cellular composition of BAL fluid in patients with severe SARS-CoV-2 pneumonia.** 18 BAL fluid samples were analyzed using a published flow cytometry protocol. Because of the small numbers in the study, we decided a priori to analyze all samples treated with Auxora or placebo together, irrespective of dosing schedule. G1 = group 1 (72 hours); G2 = group 2 (96 hours); P = placebo.

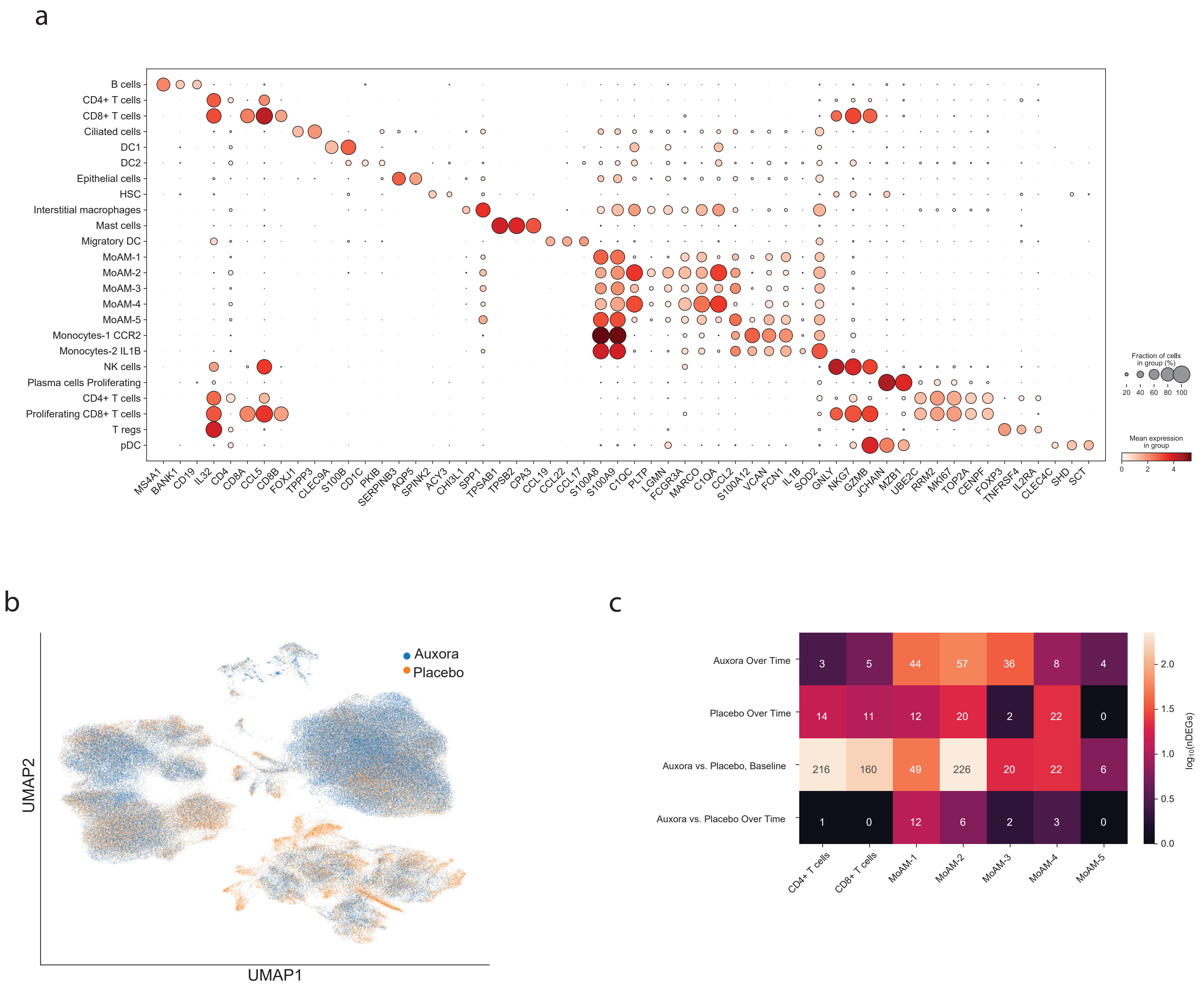


**Supplementary Figure 3: Single cell analysis of BAL fluid samples collected before and after treatment with Auxora or placebo. a**, Dot plot illustrating expression of marker genes for scRNA-seq cluster annotations. Cell phenotypes are listed on the y-axis and genes (features) are listed along the x-axis. Dot size reflects the percentage of cells in a cluster expressing each gene; dot color reflects expression level. **b**, UMAP plot illustrating integrated analysis of BAL cells isolated from patients with severe SARS-CoV-2 pneumonia (n=9) and labeled for placebo or Auxora treatments. **c**, cell abundances before and after administration of Auxora measured using scRNA-sequencing. Neutrophils were excluded by our cryopreservation procedure and their abundance was estimated from flow cytometry data.

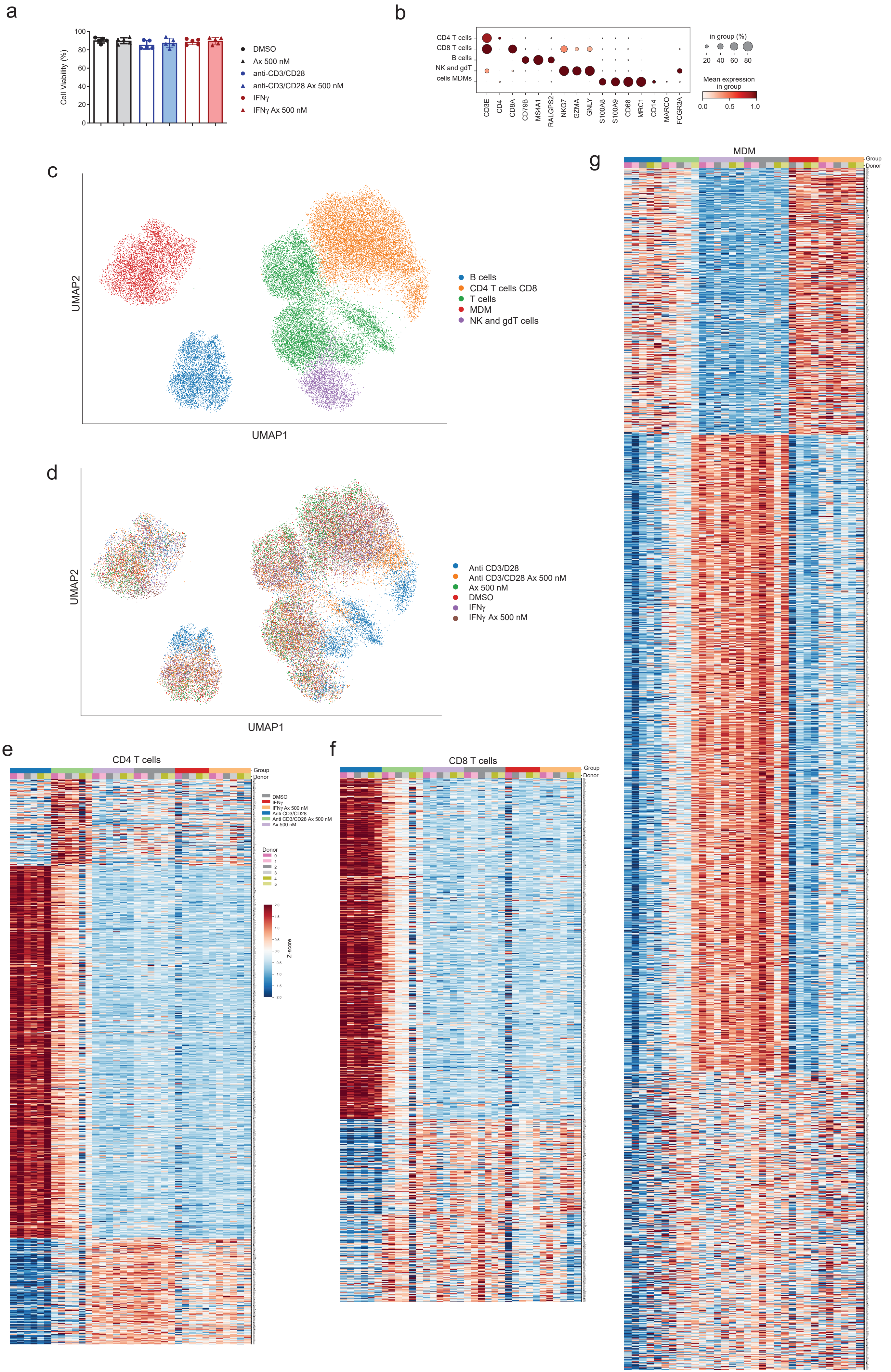
a
Cell Viability (%)
100
80
60
40
20
0
DMSO
Ax 500 nM
anti-CD3/CD28
anti-CD3/CD28 Ax 500 nM
IFNγ
IFNγ Ax 500 nM
b
CD4 T cells
CD8 T cells
B cells
NK and gdT
cells MDMs
CD3E
CD4
CD8A
CD79B
MS4A1
RALGPS2
NKG7
GZMA
GNLY
S100A8
S100A9
CD68
MRC1
CD14
MARCO
FCGR3A
in group (%)
20 40 60 80
Mean expression in group
0.0 0.5 1.0
c
UMAP2
UMAP1
B cells
CD4 T cells CD8
T cells
MDM
NK and gdT cells
d
UMAP2
UMAP1
Anti CD3/D28
Anti CD3/CD28 Ax 500 nM
Ax 500 nM
DMSO
IFNγ
IFNγ Ax 500 nM
e
CD4 T cells
Group
Donor
DMSO
IFNγ
IFNγ Ax 500 nM
Anti CD3/CD28
Anti CD3/CD28 Ax 500 nM
Ax 500 nM
Donor
0
1
2
3
4
5
Z-score
f
CD8 T cells
Group
Donor
g
MDM
Group
Donor

__Supplementary Figure 4:__ **Auxora reverses gene transcriptional changes in PBMCs stimulated with anti-CD3/CD28 antibodies but not in PBMCs stimulated with IFN-γ.** PBMCs were plated for 1 day and treated with DMSO or Auxora (Ax) 500 nM for 2 h. Then, cells were exposed to anti-CD3/ CD28 antibodies, IFN-γ or media as control. scRNAseq was performed after 24 h. **a**, Cell viability measured using a Cellometer K2. **b**, Dot plot illustrates expression of marker genes used to annotate scRNA-seq data. Dot size reflects the percentage of cells in a cluster expressing each gene; dot color reflects expression level. and cells induced by the different treatments. **c** and **d**, UMAP plot showing integrated analysis of scRNA-seq data from cells derived from cultured PBMCs. N=5 subjects. Heat maps of gene expression from **e**, CD4 T cells; **f**, CD8 T cells; and **g**, MDM.

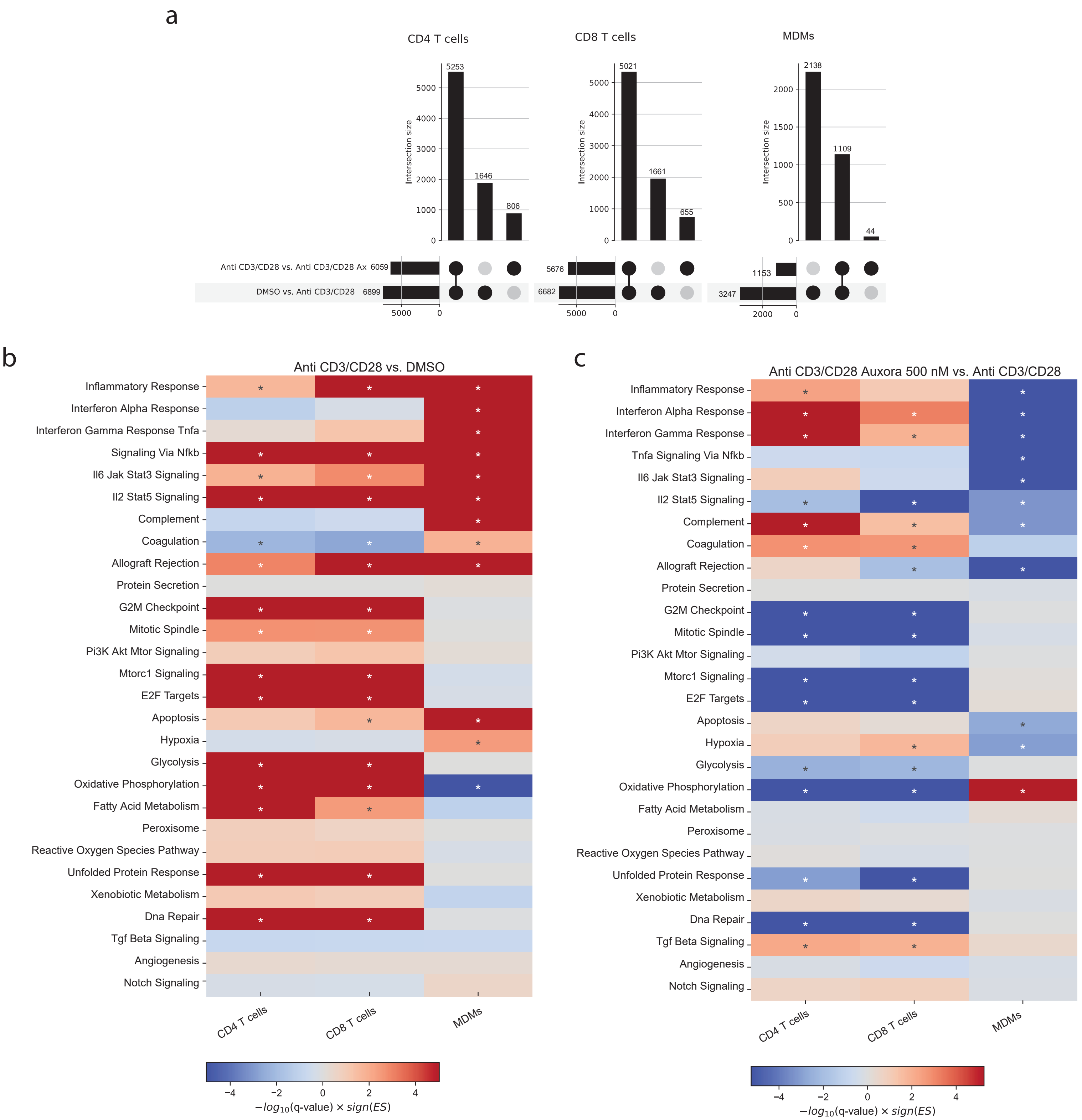


**Supplementary Figure 5: Auxora breaks T cell-macrophage circuits and prevents macrophage activation by acting on T cells.** PBMCs were plated for 1 day and treated with DMSO or Auxora (Ax) 500 nM for 2 h. Then, cells were exposed to anti-CD3/CD28 or media as control. scRNA-seq was performed after 24 h. **a**, Upset plots showing DEGs by T cell activation and by the effect of Auxora. **b**, Heatmap illustrating enrichment for 50 hallmark gene sets from MSigDB between anti-CD3/CD28 vs DMSO in CD4 T cells, CD8 T cells, and MDM. **c**, Heatmap illustrating enrichment for 50 hallmark gene sets from MSigDB between IFN-γ vs DMSO in CD4 T cells, CD8 T cells, and MDM. Significant enrichments (q-value < 0.05) are indicated by asterisks.

a

b

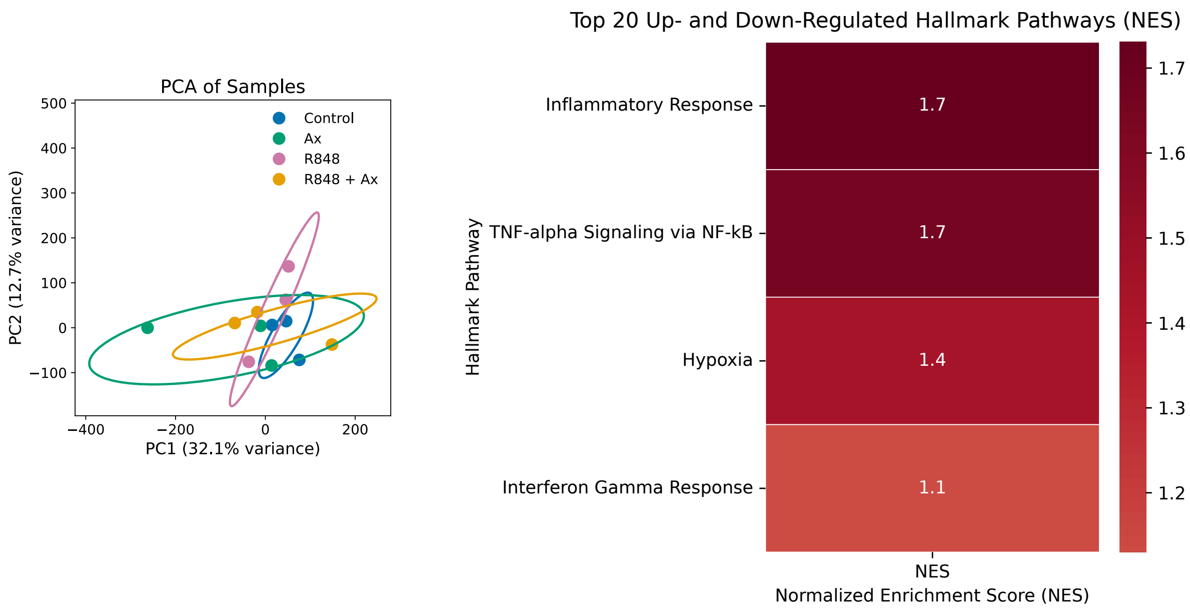


**<u>Supplementary Figure 6:</u> Auxora does not reverse transcriptional programs induced by TLR agonists in MDM.** MDMs were treated with the TLR4 agonist LPS (10 ng/ml) or the TRL7/8 agonist R848 (1 μM) in the presence of DMSO or Auxora 500 nM for 2 h and harvested for bulk RNA-seq. **a**, Principal-component analysis (PCA) was performed on differentially expressed genes identified from a generalized linear model to perform an ANOVA-like test for differential expression between any conditions in the dataset (FDR q < 0.05). **b**, Pathways activated by R848.

| Characteristic | Subcategories | Number (%) or median [Q1,Q3] |
| --- | --- | --- |
| n | | 9 |
| Age, median [Q1,Q3] | | 55.0 [50.0,64.0] |
| Sex at Birth, n (%) | Female | 3 (33.3) |
| | Male | 6 (66.7) |
| Ethnicity, n (%) | Hispanic | 6 (66.7) |
| | Non-Hispanic | 3 (33.3) |
| Race, n (%) | Black or African American | 1 (11.1) |
| | Other | 1 (11.1) |
| | Unknown or Not Reported | 3 (33.3) |
| | White or Caucasian | 4 (44.4) |
| BMI, median [Q1,Q3] | | 30.1 [27.7,35.4] |
| Co-enrolled in SCRIPT study, n (%) | No | 2 (22.2) |
| | Yes | 7 (77.8) |
| **Comorbidities** | | |
| Diabetes, n (%) | Unknown | 6 (66.7) |
| | Yes | 3 (33.3) |
| Hypertension, n (%) | Unknown | 6 (66.7) |
| | Yes | 3 (33.3) |
| Smoker, n (%) | Former smoker | 1 (11.1) |
| | Missing or N/A | 4 (44.4) |
| | Never smoked | 4 (44.4) |
| Asthma, n (%) | Unknown | 8 (88.9) |
| | Yes | 1 (11.1) |
| Chronic obstructive pulmonary disease, n (%) | No | 1 (11.1) |
| | Unknown | 8 (88.9) |
| Coronary artery disease, n (%) | Unknown | 9 (100.0) |
| Congestive heart failure, n (%) | Unknown | 9 (100.0) |
| **Medication information** | | |
| Received azithromycin, n (%) | No | 3 (33.3) |
| | Yes | 6 (66.7) |
| Ongoing azithromycin at randomization, n (%) | No | 3 (33.3) |
| | Yes | 3 (33.3) |
| Received remdesivir, n (%) | No | 2 (22.2) |
| | Yes | 7 (77.8) |
| Ongoing remdesivir at randomization, n (%) | No | 3 (33.3) |
| | Yes | 4 (44.4) |
| Received Corticosteroids, n (%) | No | 2 (22.2) |
| | Yes | 7 (77.8) |
| Received Dexamethasone, n (%) | Yes | 7 (77.8) |
| | No | 2 (22.2) |
| Ongoing corticosteroids at randomization, n (%) | Yes | 7 (77.8) |

**<u>Supplementary Table 1:</u> Demographic and clinical characteristics of the study population.** This table presents general participant demographics, including age, sex, ethnicity, and race. Clinical variables include comorbidities, as well as medications received, and whether these were ongoing at time of study drug randomization.
