## Supplementary material for "The CRAC channel inhibitor Auxora interrupts inflammatory circuits between alveolar macrophages and T cells in patients with viral pneumonia": Methods

#### Human subjects

All research involving human participants was approved by the Institutional Review Board of Northwestern University. Volunteers were enrolled in study STU00214054 and study STU00204868 at Northwestern University. We enrolled patients in a single-center, single-blind placebo-controlled ascending-dose phase 2 clinical trial to generate pharmacokinetic (PK) and pharmacodynamic (PD) data in critically ill, mechanically ventilated patients with laboratory-confirmed SARS-CoV-2 pneumonia receiving Auxora or placebo. Inclusion criteria were laboratory-confirmed SARS-CoV-2 pneumonia, moderate acute respiratory distress syndrome (PEEP > 5 cmH2O, $PaO_2/FiO_2 < 200$, and no evidence of volume overload or heart failure as the primary etiology of respiratory failure). The main exclusion criteria included mechanical ventilation > 7 days, septic shock, organ or hematologic transplant, and QTcF interval > 440 msecs. Based on our prior study[1], the primary objective was to assess the PD response of bronchoalveolar lavage (BAL) fluid T cell and monocyte and macrophage subsets and cytokine/chemokine levels to various doses of Auxora. Secondary objectives were 1) to assess safety and tolerability and 2) to determine the serum and BAL fluid PK profile of Auxora in mechanically ventilated patients with severe SARS-CoV-2 pneumonia. The trial was designed to enroll 4 patients each into three cohorts in a 3:1 drug:placebo ratio. The first cohort received Auxora or placebo once daily for 3 days, the second cohort received Auxora or placebo once daily for 4 days, and the third cohort received Auxora as a continuous infusion for 5 days. In cohorts 1 and 2, the dose of Auxora was 2.0 mg/kg (1.25 mL/kg) administered as a 4-hour infusion at 0 hour, and then 1.6 mg/kg (1 mL/kg) at 24, 48 (cohort 1), and 72 hours (cohort 2) from the start of the first infusion. In cohort 3, after an initial infusion of 2.0 mg/kg Auxora over 4 hours, the patient received continuous infusion of 1.6 mg/kg over 24 hours for 96 hours. For all cohorts, a BAL procedure was performed within 12 hours before the first dose of Auxora or placebo and 24 hours after the last dose of Auxora or placebo. One consented patient developed a prolonged QT interval prior to infusion and was excluded; another patient was recruited to fill the cohort. The trial, which coincided with the end of the COVID-19 pandemic, was stopped after only one patient who received drug was enrolled in the third cohort due to low enrollment.

#### Bronchoscopy and BAL

Bronchoscopic BAL was performed as previously described[1].

#### SARS-CoV-2 viral titers

SARS-CoV-2 viral titers were estimated from the RT-qPCR cycle threshold of BAL fluid.

#### Multiplexed cytokine assay processing and analysis

Processing and high-level analysis were performed as previously described[2]. Briefly, raw MFI values, bead counts, and standard concentrations were first stripped from the data output from Eve Technologies (Calgary, Alberta, Canada). MFI measurements with fewer than 50 bead counts were discarded. Standard curves for each cytokine were then fit for each assay run using self-starting 5-parameter logistic (5PL) models using drc 3.2-0. Cutoffs for curves with low predictive value were then determined empirically using histograms' MFI values versus standard concentrations to identify a bimodal distribution cutoff. For in-house assays, all values calculated using standard curves with MFI < 50 at 100 pg/mL were discarded. For Eve Technologies assays, all values calculated using standard curves with MFI < 50 at 10 pg/mL were discarded. Experimental values for each cytokine were then predicted using the ED function in drc with "absolute" value prediction. In rare cases where a 5PL model could not be fit for an individual cytokine assay combination, these values were excluded. Values below the lower asymptote of the model were set to a concentration of 0 pg/mL. Values above the upper asymptote were set to the value of the upper asymptote. Technical replicates (including those across assays) were collapsed by mean with NA values excluded. Analytes showing poor dynamic range were excluded from further analysis. To identify cytokines responding to Auxora, linear mixed models using a paired design, considering treatment status (before or after treatment), treatment group (Auxora or placebo) and patient were applied for each cytokine with the formula [cytokine] ~ treated * treatment_group + (1|patient) using lme4 1.1-30. P-values were then extracted for the term "treatedTRUE:treatment_groupDrug".

**BAL sample processing for scRNA-seq using "no FACS" protocol**

BALF samples were filtered through a 70-µm cell strainer, pelleted by centrifugation at 400 rcf for 10 min at 4°C, followed by hypotonic erythrocyte lysis using 1 ml of PharmLyse solution (BD) for 2 min, followed by another wash with 10 ml of 0.5% BSA in PBS and centrifugation. Cell pellet was resuspended in 0.5% BSA in PBS at the final concentration 1000 cells/ul. Cell concentration and viability were determined using K2 Cellometer (Nexcelom) with AO/PI reagent and cells were loaded on 10x Genomics Chip A with Chromium Single Cell 3′ V2 gel beads and reagents (10x Genomics) aiming to capture 5,000–7,000 cells per library. Libraries were prepared according to the manufacturer's protocol (10x Genomics, CG000052_RevB). BAL sample processing for scRNA-seq using "FACS" protocol: BAL fluid samples were filtered through a 70-µm cell strainer, pelleted by centrifugation at 400 relative centrifugal force (rcf) for 10 min at 4 °C, followed by hypotonic lysis of red blood cells with 2 ml of PharmLyse (BD Biosciences) reagent for 2 min. Lysis was stopped by adding 13 ml of MACS buffer (Miltenyi Biotech). Cells were pelleted again and resuspended in 100 µl of a 1:10 dilution of Human TruStain FcX (BioLegend) in MACS buffer and a 10-µl aliquot was taken for counting using a K2 Cellometer (Nexcelom) with Acridine orange (AO)/Propidium iodide (PI) reagent. The cell suspension volume was adjusted so the concentration of cells was always $< 5 \times 10^7$ cells/ ml and the fluorophore-conjugated antibody cocktail was added in a 1:1 ratio. The following antibodies were used (antigen, clone, fluorochrome, manufacturer, catalog no., final dilution): CD4, RPA-T4, BUV395, BD, 564724, 1:40; CD19, HIB19, BUV395, BD, 740287, 1:40; CD25, 2A3, BUV737, BD, 564385, 1:20; CD56, NCAM16.2, BUV737, BD, 612766, 1:20; HLA-DR, L243, eFluor450, Thermo Fisher Scientific, 48-9952-42, 1:40; CD45, HI30, BV510, BioLegend, 304036, 1:20; CD15, HI98, BV786, BD, 563838, 1:20; CD3, SK7, PE, Thermo Fisher Scientific, 12-0036-42, 1:20; CD127, HIL-7R, PECF594, BD, 562397, 1:20; CD206, 19.2, PECy7, Thermo Fisher Scientific, 25-2069-42, 1:40; CD8, SK1, APC, BioLegend, 344721, 1:40; CD14, M5E2, APC, BioLegend, 301808, 1:40; and EpCAM, 9C4, APC, BioLegend, 324208, 1:40. After incubation at 4 °C for 30 min, cells were washed with 5 ml of MACS buffer, pelleted by centrifugation and resuspended in 500 µl of MACS buffer with 2 µl of SYTOX Green viability dye (Thermo Fisher Scientific). Cells were sorted on a FACS Aria III SORP instrument using a 100-µm nozzle. Cells were sorted into 300 µl of 2% bovine serum albumin (BSA) in Dulbecco's phosphate-buffered saline (DPBS) and immediately after sorting pelleted by centrifugation at 400 rcf for 5 min at 4 °C, resuspended in 0.5 BSA in DPBS to 1,000 cells/ µl concentration. Concentration was confirmed using a K2 Cellometer (Nexcelom) with AO/PI reagent using the 'Immune cells low RBC' program with default settings and cells were immediately used for scRNA-seq. Cells were loaded on 10x Genomics Chip A with Chromium Single Cell 3′ V2 gel beads and reagents or 10x Genomics Chip B with Chromium Single Cell 3′

V3 gel beads and reagents (10x Genomics) aiming to capture 5,000–7,000 cells per library. Libraries were prepared according to the manufacturer's protocol (10x Genomics, CG000052_RevB or CG000183_RevB).

Analysis of the flow cytometry data was performed using FlowJo v.10.7.1. using a sequential gating strategy reported in our previous publications. A fraction of cells that was not definitively resolved by our panel was labeled 'others'. Relative cell-type abundance was calculated as a percentage of all singlets/live/CD45+ cells. Statistical methods: No statistical method was used to predetermine sample size. The experiments were not randomized. The Investigators were not blinded to allocation during experiments and outcome assessment. All statistics in the manuscript are reported as specified in the figure legends. When multiple hypothesis tests were performed, the false discovery rate (FDR) was controlled using the procedure of Benjamini and Hochberg. A significance level of 0.05 was used for all tests, unless indicated otherwise.

**PBMC isolation**

PBMC samples were isolated from blood of 5 healthy individuals and collected in Vacutainer EDTA tubes (generic laboratory supplier) using SepMate-50 tubes (STEMCELL Technologies) following manufacturer's protocol. Blood samples were diluted 1:1 with PBS/ 2% FBS. The diluted blood was layered on top of a 15 ml Lymphoprep™ (STEMCELL Technologies) layer in the SepMate tube. The SepMate tubes were centrifuged at 1,200g for 10 min at room temperature. After centrifuging, the top plasma layer was removed as much as possible without disturbing the PBMC layer. The cells were poured from the SepMate tube into a 50 ml conical tube. The tubes containing cells were filled up to 50 ml with cold wash buffer (PBS with 2% FBS) and mixed by inverting. The tubes were centrifuged at 300g for 10 min at room temperature. After centrifuging, the supernatant was removed without disturbing the cell pellet. After resuspending the pellet with cold wash buffer, the cells were counted using the Nexcelom K2 Cellometer C automated cell counter with AO/ PI reagent. The tubes were again centrifuged at 300g for 10 min with brake set to 5 at room temperature. The supernatant was removed without disturbing the cell pellet. Aliquots containing $1.5 \times 10^6$ PBMCs in 200 µL Bambanker Cell Freezing Media were stored into a −80 °C freezer.

**Preparation of Auxora for experimental use**

The Orai1 inhibitor compound is a dry powder manufactured and provided by CalciMedica. Auxora was dissolved in DMSO preparing a 100 mM stock solution and was used in 50 and 500 nM working concentration on cell cultures.

**PBMC stimulation**

PBMCs ($1 \times 10^5$ cells/ well) were plated in RPMI (ATCC) with 10% heat-inactivated FBS with 100 U/ml Penicillin/Streptomycin in Ultra-Low Cluster 96 well plates (Costar) for 1 day. Then, PBMCs were treated with DMSO or Auxora at 50 nM or 500 nM for 2 h and T cells were exposed to ImmunoCult™ Human CD3/CD28/CD2 T Cell Activator (25 µl/ml, STEMCELL Technologies), IFNγ (50 ng/mL) or media as control. scRNAseq was performed after 24 h.

**Differentiation and activation of human monocytes derived macrophages (MDM) with LPS and R484**

Monocytes were isolated from PBMCs using the EasySep™ Human Monocyte Isolation Kit (STEMCELL Technologies). Briefly, this kit is designed to isolate $CD14^+$ $CD16^-$ monocytes from frozen PBMCs samples by negative selection of unwanted cells and platelets with antibody complexes and magnetic particles. Then, these monocytes were differentiated to MDMs using ImmunoCult™-SF Macrophage Medium plus 5 µg/mL human recombinant M-CSF (both from STEMCELL Technologies) for 4 days and 10 ng/mL LPS and 50 ng/mL IFN-γ (STEMCELL Technologies) for additional 2 days. Then, MDMs were incubated for 2 h with Auxora (500 nM) or DMSO and then activated with the TLR7/TLR8 ligand R848 (1 µM, InvivoGen). RNA was extracted after 24 h for bulk RNAseq.

**PBMC single-cell RNA sequencing**

PBMCs single-cell suspensions were prepared as described earlier. Cells were counted using a Cellometer K2 with nucleic acid binding dyes AO to calculate total number of nucleated cells and PI to count dead cells; cell viability exceeded 85%. All manipulations were performed using wide-bore tips (Axygen; Corning). Single-cell 3′ RNA sequencing libraries were prepared using Chromium Single Cell v2 Reagent Kit and Controller (10X Genomics, Pleasanton, CA, USA). Libraries were assessed for quality (TapeStation 4200; Agilent, Santa Clara, CA, USA) and then sequenced on an HiSeq 4000 instrument (Illumina, San Diego, CA, USA). Data was processed as shown above.

**Single-cell RNA-seq analysis**

PBMC data were processed using Cell Ranger 1.1.0 (10x Genomics) and reads were mapped to the GRCh38 reference genome (version refdata-gex-GRCh38-2020-A, 10x Genomics). Per-sample doublet detection was performed with Scrublet[3]. Data were processed using Scanpy 1.9.8, and multisample integration was performed with scvi-tools 1.1.1. An initial scVI model was constructed on 2000 HVGs with the hyperparameters n_layers = 2, n_hidden = 256, dropout_rate = 0.2, and n_latent = 10, and was trained with max_epochs = 400 and early_stopping = True. Default hyperparameters and settings were used otherwise. Leiden clustering was performed on the integrated object, and clusters characterized by low number of detected genes and transcripts, high percentage of mitochondrial genes, or co-expression of lineage-specific markers from distinct cell types (indicating doublets) were excluded. The remaining cells were re-integrated with scVI using the same hyperparameters on 1000 HVGs, with T cell receptor genes (gene symbols starting with TR) excluded prior to HVG selection to prevent clonotype-specific expression from driving the embedding of T cell populations. Leiden clustering was performed on the second integration with a resolution of 1. Cell types were identified by marker genes, computed using the sc.tl.rank_genes_groups function with default settings. Donor identity for each cell was assigned using Souporcell[4] with k = 5 expected genotype clusters per sequencing library, and cells classified as doublets or unassigned were excluded from downstream analyses.

BAL data were processed using Cell Ranger 3.1.0 (10x Genomics) and reads were mapped to the GRCh38.84 reference genome (10x Genomics Cell Ranger Human 1.2.0 GRCh38 reference). Data was processed using Scanpy 1.10.4, and multisample integration was performed with scvi-tools 1.3.0. The scVI model was constructed on 2000 HVGs with the hyperparameters n_layers = 2, dropout_rate = 0.2, and n_latent = 10, and were trained using the settings max_epochs = 400, check_val_every_n_epoch = 2, and early_stopping = True. Default hyperparameters and settings were used otherwise. An initial round of Leiden clustering using the function sc.tl.leiden was performed on the integrated BAL object with a resolution of 1. Clusters characterized by low number of detected genes and transcripts and high percentage of mitochondrial genes were removed. Clusters containing doublets were identified as clusters simultaneously expressing lineage-specific marker genes (for example, C1QA for macrophages and CD3G for T cells) and excluded. Cell types were identified by marker genes, computed using the sc.tl.rank_genes_groups function with default settings.

For more details, see code. Differential abundance analysis was performed on fractions of identified cell clusters in each PBMC/BAL sample out of the whole sample, and the comparison was done with Mann-Whitney U tests.

**Pseudobulk gene expression profiles**

For several analyses of gene expression, including MOFA, PCA, differential gene expression analysis, and gene set variation analysis, we summarized gene expression for each sample and each annotated cell type from our scRNA-seq data. We required at least 50 cells to create a pseudobulk sample for a given sample and cell type. We summed integer counts for all cells across all genes, and saved this matrix for all cell types. We excluded several gene categories from the pseudobulk count matrices: mitochondrial genes, SARS-CoV-2 viral transcripts, ribosomal protein-coding genes (RPL and RPS families), and unannotated transcripts lacking canonical HGNC symbols (i.e., genes identified only by Ensembl or NCBI/RefSeq accession identifiers). These categories were excluded a priori because they reflect technical artifacts, viral rather than host signal, or transcripts without interpretable biological annotation, as we sought to elucidate established biological pathways as well as credential protein based biomarkers.

**Differential gene expression analysis**

To identify genes that are expressed differently between groups of samples, we used a modified pseudobulk method. This method demonstrates better consistency with the bulk approach in benchmarking studies[5], compared to single cell based methods for testing of differentially expressed genes. Additionally, it avoids the issue of inflated p-values. We used pseudobulk gene count matrices as described above and constructed a DESeq2 object using either all BAL samples or PBMC samples. For the DESeq2 model for the BAL analysis, we used ~ group + group:pair_id + group:timepoint, which models paired pre- and post-treatment BAL samples within each treatment group and tests whether the timepoint response differs between Auxora and Placebo, and we used fitType local. Differentially expressed genes were obtained from the Auxora time point 2 vs. Placebo time point 2 contrast (Wald test, $\alpha = 0.05$). For the DESeq2 model for the PBMC analysis, we used ~ pair_id + group, where pair_id controls for baseline inter-donor variability and group encodes the stimulation/Auxora condition, and we used fitType local. Differentially expressed genes were obtained from the Ax 500 nM vs. DMSO, Anti CD3/CD28 vs. DMSO, Anti CD3/CD28 Ax 500 nM vs. Anti CD3/CD28, IFNy vs. DMSO, and IFNy Auxora 500 nM vs IFNy contrasts (Wald test, $\alpha = 0.05$).

**Wide-field fura-2 $Ca^{+2}$ imaging**

T cells or MDMs incubated on poly-d-lysine–coated glass-bottom dishes were loaded with Fura-2 by incubating cells in 2 µM Fura-2-AM (Invitrogen, F1221) in growth medium for 30 min at 37°C. Fura-2–containing medium was washed off, and cells were incubated for an additional 10 min before imaging. All experiments were performed at room temperature. Single-cell $[Ca^{2+}]_i$ measurements were performed as described previously[6,7]. Image acquisition and analysis were performed using SlideBook (Denver, CO). Dishes were mounted on the stage on Olympus IX71 inverted microscope, and images were acquired every 6 s at excitation wavelengths of 340 and 380 nm and an emission wavelength of 510 nm. For data analysis, regions of interest (ROIs) were drawn around single cells, background was subtracted, and F340/F380 ratios were calculated for each time point.

$[Ca^{2+}]i$ was estimated from F340/F380 ratio using the standard equation: $[Ca^{2+}]i = \beta Kd (R - Rmin)/(Rmax - R)$, where R is the F340/F380 fluorescence ratio and values of Rmin and Rmax were determined from an in vitro calibration of Fura-2 pentapotassium salt. β was determined from the F340/F380 ratio at 380 nm, and Kd is the apparent dissociation constant of Fura-2 binding to $Ca^{2+}$ (135 nM). For each cell, the rate of SOCE ($\Delta[Ca^{2+}]i/\Delta t$) was calculated from the slope of a line fitted to three points (18 s) after the readdition of 2 mM Ca2+o. Baseline $[Ca^{2+}]_i$ was calculated by averaging $[Ca^{2+}]_i$ values over 2-min baseline for each experiment. Peak $Ca^{2+}$ was calculated by determining the maximum $Ca^{2+}$ concentration when 2 Ca was added back after store depletion with thapsigargin (Tg). The standard Ringer's solution used for these experiments contained the following: 155 mM NaCl, 4.5 mM KCl, 10 mM d-glucose, 5 mM Hepes, 1 mM $MgCl_2$, and 2 mM $CaCl_2$. The $Ca^{2+}$-free Ringer's solution was similar to the above solution except that it contained 3 mM $MgCl_2$ and 1 mM EGTA (Sigma-Aldrich) with no added $CaCl_2$. pH was adjusted to 7.4 with 1 M NaOH. The stock solution of Tg was dissolved in dimethyl sulfoxide (DMSO) and used at the indicated concentration.

### Figure and manuscript preparation

Plotting was performed in Python using matplotlib and seaborn. Specific details are below and in the included code. Figures were assembled in Adobe Illustrator.

### Data availability

Single-cell RNA-seq counts tables and integrated objects are available through the Gene Expression Omnibus with accession number GSE330843.

### Code availability

All code used for analysis can be found at https://github.com/NUPulmonary/Guggilla_Matsuda_Auxora_2026. ScRNA-seq data can be explored via UCSC data browsers at https://sqlifts.fsm.northwestern.edu/internal/auxora/?ds=pbmc_integrated_0424 and at https://sqlifts.fsm.northwestern.edu/internal/auxora/?ds=2025-09-30_Auxora_Human.

### *References*


1 Grant, R. A. *et al.* Circuits between infected macrophages and T cells in SARS-CoV-2 pneumonia. *Nature* **590**, 635-641 (2021). https://doi.org/10.1038/s41586-020-03148-w

2 Grant, R. A. *et al.* Prolonged exposure to lung-derived cytokines is associated with activation of microglia in patients with COVID-19. *JCI Insight* **9** (2024). https://doi.org/10.1172/jci.insight.178859

3 Wolock, S. L., Lopez, R. & Klein, A. M. Scrublet: Computational Identification of Cell Doublets in Single-Cell Transcriptomic Data. *Cell Syst* **8**, 281-291 e289 (2019). https://doi.org/10.1016/j.cels.2018.11.005

4 Heaton, H. *et al.* Souporcell: robust clustering of single-cell RNA-seq data by genotype without reference genotypes. *Nat Methods* **17**, 615-620 (2020). https://doi.org/10.1038/s41592-020-0820-1

5 Squair, J. W. *et al.* Confronting false discoveries in single-cell differential expression. *Nat Commun* **12**, 5692 (2021). https://doi.org/10.1038/s41467-021-25960-2

6 Feske, S. *et al.* A mutation in Orai1 causes immune deficiency by abrogating CRAC channel function. *Nature* **441**, 179-185 (2006). https://doi.org/10.1038/nature04702

7 Soberanes, S. *et al.* Metformin Targets Mitochondrial Electron Transport to Reduce Air-Pollution-Induced Thrombosis. *Cell Metab* **29**, 503 (2019). https://doi.org/10.1016/j.cmet.2018.12.001
